# Longitudinal analysis of the prevalence of minor *Plasmodium* spp. in the reservoir of asymptomatic infections through sequential interventions in Northern Sahelian Ghana

**DOI:** 10.1101/2025.08.15.25333797

**Authors:** Cecilia A. Rios-Teran, Kathryn E. Tiedje, Oscar Bangre, Samantha L. Deed, Dionne C. Argyropoulos, Kwadwo A. Koram, Abraham R. Oduro, Patrick O. Ansah, Karen P. Day

## Abstract

Current interventions targeting malaria control in sub-Saharan Africa (SSA) are focused on *Plasmodium falciparum*, the most prevalent species infecting humans. Despite renewed efforts for malaria elimination in SSA, little attention has been paid to the neglected parasites *P. malariae* and *P. ovale* spp. and the impact of interventions like long-lasting insecticidal nets (LLINs), indoor residual spraying (IRS) with non-pyrethroid insecticides, and/or seasonal malaria chemoprevention (SMC) on these minor *Plasmodium* spp. To address this research gap, this study was undertaken to assess the efficacy of two sequential interventions, IRS and SMC combined with LLINs, on minor-*Plasmodium* spp. infections in Bongo District, an area characterized by high seasonal transmission in the Northern Sahelian belt of Ghana. Using an interrupted time-series study, five age-stratified surveys, each of ∼2,000 participants, were undertaken at the end of the wet seasons between 2012 and 2022. Across this 10-year study period, infections with *P. malariae* and *P. ovale* spp. were detected using a species-specific PCR targeting the *18S rRNA* gene. In 2015, following IRS, the prevalence of the minor *Plasmodium* spp. declined in all ages from baseline in 2012, with participants being significantly less likely to be infected with *P. malariae* (13.7% vs 1.4%) and *P. ovale* spp. (5.7% vs 0.4%). Despite this decline, in 2017, 32 months after IRS was discontinued and SMC was introduced, the prevalence of *P. malariae* (2.9%) and *P. ovale* spp. (4.0%) rebounded 2- and 10-fold, respectively. This rebound in the minor species was observed in all age groups, except for the younger children (< 5 years) targeted by SMC. Finally, when we examined this population in 2020 and 2022 after sustained deployment of SMC, the prevalence of *P. malariae* continued to increase (7.4% and 5.8%), while the prevalence of *P. ovale* spp. declined (2.6% and 1.3%). Results show that both IRS and SMC were effective not only against *P. falciparum* but also reduced the prevalence of *P. malariae* and *P. ovale* spp. in Bongo District. Going forward, molecular diagnostics will be critical to identify changes in the submicroscopic reservoir of the minor *Plasmodium* spp. found in adolescents and adults and to achieve malaria elimination in this region of Ghana.

**AUTHOR SUMMARY:** To achieve malaria elimination, the use of molecular diagnostics for surveillance of the *Plasmodium* spp. reservoir is essential. Such diagnostics are critical to detect hidden reservoirs of submicroscopic minor-species infections with *P. malariae* and *P. ovale* spp. Here we examine the impact of two sequential malaria control interventions, indoor residual spraying (IRS) with non-pyrethroid insecticides and seasonal malaria chemoprevention (SMC) with sulfadoxine-pyrimethamine plus amodiaquine administered to all children < 5 years, combined with long-lasting insecticidal nets (LLINs) impregnated with pyrethroids to reduce the reservoir of *Plasmodium* spp. infections where *P. falciparum* is dominant and the minor species, *P. malariae* and *P. ovale* spp., were also found. This interrupted time-series study, undertaken over 10 years (2012-2022) in Bongo District, located in the Northern Sahelian belt of Ghana, identified that IRS and SMC reduced the prevalence of all *Plasmodium* spp. IRS significantly reduced the prevalence of single- and mixed-*Plasmodium* spp. infections across all age groups. However, following its discontinuation a rebound was observed for *P. falciparum* and minor-species infections, especially in adolescents and adults. Although SMC maintained a near-zero prevalence of all *Plasmodium* spp. infections, its impact was limited to the target age group (< 5 years). Furthermore, this study showed that following the discontinuation of IRS, the older children, adolescents, and adults, not eligible for SMC, continued to harbour *Plasmodium* spp. infections, including minor species, thus sustaining the reservoir. These results show that by combining vector control inventions as well as targeting all age groups with chemoprevention, high-burden countries in sub-Saharan Africa, including Ghana, could bring the *Plasmodium* spp. reservoir closer to elimination more rapidly.

## INTRODUCTION

Since the early 2000s much of sub-Saharan Africa (SSA) has experienced scale-up of malaria control interventions, including long-lasting insecticidal nets (LLINs), indoor residual spraying (IRS), and the treatment of clinical malaria with artemisinin-based combination therapies (ACTs) [1]. These interventions have successfully reduced childhood morbidity and mortality due to malaria but very little is known about the impact of these interventions on the reservoir of asymptomatic *Plasmodium* spp. infections that sustain transmission to anopheline mosquitoes, especially in high transmission.

To achieve malaria elimination in high-burden countries in SSA the challenge remains to reduce the reservoir of these asymptomatic infections to zero. Thus, it is critical to understand the composition, population size, and age distribution of this *Plasmodium* spp. reservoir in humans and the parasitological changes that occur following the implementation of the abovementioned control interventions. To date, there have been limited studies of this reservoir using sensitive molecular diagnostics to investigate the prevalence of all *Plasmodium* spp infecting humans. Contemporary studies in SSA have largely focused on the dominant species, *Plasmodium falciparum*, with few investigations of the minor species, *P. malariae*, *P. ovale* spp. And *P. vivax*, that are often neglected (e.g., [2–7]). Furthermore, there have only been a limited number of studies from high-transmission settings that have monitored the impact of the abovementioned interventions on the prevalence of *P. falciparum* in the reservoir of infection (e.g., [8–12]). None have concurrently evaluated the minor-*Plasmodium* spp. infections with sensitive molecular diagnostics Typical of SSA, Ghana has intensified its national malaria control efforts through the implementation of various intervention strategies (i.e., LLINs, IRS, seasonal malaria chemoprevention (SMC), etc.), across the country’s different epidemiological or ecological zones [13–16]. IRS, in combination with LLINs, has been deployed across much of northern Ghana, specifically, in the Upper West Region since 2011, the Northern Region since 2012, and the Upper East Region since 2013 [16–18]. However, its implementation has not been continuous across all districts. The timing and geographic scale-up of IRS in these regions have been associated with positive impacts on malaria control, as evidenced by reductions in the incidence of confirmed clinical cases [17]. In the Upper West Region, where IRS has been combined with SMC administered to all children between 3-59 months (< 5 years) since 2015, data show a sustained reduction in clinical malaria incidence [17]. Despite these successes, the impact of these interventions on the reservoir of *Plasmodium* spp. infection, remains unknown. To address this parasitological research gap, the Malaria Reservoir Study (MRS) was established in Bongo District in the Upper East Region to follow changes in the parasite population in humans from a baseline in 2012 through the implementation of sequential interventions out to 2022 [12,19,20]. This is the only such longitudinal study of the reservoir of *Plasmodium* spp. infection in all ages in West Africa with the benefit of a baseline, prior to implementation of IRS and SMC.

In Bongo District, located in the Northern Sahelian belt of Ghana, no studies have focused exclusively on characterising minor *Plasmodium* spp. in the asymptomatic reservoir using molecular diagnostics. While not the primary objective, the MRS previously detected *P. malariae* and *P. ovale* spp. in this district using a species-specific PCR targeting the 18S ribosomal RNA gene (*18S rRNA*) [19], supporting the presence of these species and underscoring the need for a more comprehensive investigation. This reflects a broader pattern across Ghana, where relatively few studies have examined minor *Plasmodium* spp. infections in community settings across all age groups. Much of the existing evidence comes from cross-sectional studies conducted in the Eastern Region, specifically in the Kwahu-South and Akwapim South Municipal Districts [3,21]. Additional studies have identified minor *Plasmodium* spp. in specific age groups, such as asymptomatic school-aged children in the Central and Ashanti Regions [5,22,23] and asymptomatic adults in south-central Ghana, from multiple sites in the forest ecological zone [24] as well as the Ashanti Region [25]. Consequently, the prevalence of minor *Plasmodium* spp. in the asymptomatic reservoir remains largely underdetermined in northern Ghana, emphasizing the need for additional epidemiological studies.

Using an interrupted time-series study design, five age-stratified cross-sectional surveys of ∼2,000 participants per survey were undertaken at the end of the wet seasons between 2012 and 2022 to evaluate the impact of two sequential interventions, IRS and SMC, combined with widespread LLIN usage, on the prevalence and composition of the asymptomatic *Plasmodium* spp. reservoir in Bongo District, in particular the non-falciparum minor species *P. malariae*, *P. ovale* spp., and *P. vivax*.

## METHODS

### Ethical approval

This study was reviewed and approved by the ethics committees at the Navrongo Health Research Centre, Ghana (NHRC IRB-131), Noguchi Memorial Institute for Medical Research, Ghana (NMIMR-IRB CPN 089/11-12; NMIMR-IRB CPN 066/20-21), The University of Chicago, United States (IRB14-1495; IRB19-0760; IRB21-0417), New York University, United States (IRB-FY2024-8572), and The University of Melbourne, Australia (Project IDs 13433, 31586, 21649). Individual informed consent was obtained mainly in the local language (i.e., Gurene) from each participant enrolled by signature or thumbprint, accompanied by the signature of an independent witness. For children < 18 years of age, a parent or guardian provided consent. In addition, all children between the ages of 12 and 17 provided assent.

### Study site and population

This study was undertaken in Bongo District, located in the Upper East Region within the Northern Sahelian belt of Ghana, proximal to the border with Burkina Faso (Fig 1A). This district is characterized by high seasonal malaria transmission, where there is a short but intense wet season between June and October (i.e., high-transmission season) and a prolonged dry season between November and May (i.e., low-transmission season), typical of endemic regions at the southern edge of the Sahel sub-region. In Bongo District, *P. falciparum* is the dominant species, while the minor species *P. malariae* and *P. ovale* spp. are also found[19]. For this study, participants were enrolled from two broad catchment areas (i.e., Vea/Gowrie and Soe, with a sampling area of ∼60 km^2^) in Bongo District (hereinafter referred to collectively as “Bongo”) selected as they were comparable in terms of population size, age distribution, and ethnic composition but are in different agroecological zones [19]. Vea/Gowrie is located in the south of the district and is proximal to the Vea Dam and irrigation area [19,26], while Soe is situated in the north-west near the border with Burkina Faso and not located near any large bodies of water, although smaller dams for watering animals and gardening are scattered across the catchment area (Fig 1A). Details on the study site, study, population, inclusion/exclusion criteria, and data collection procedures have been previously published [12,19].

**Fig 1.**
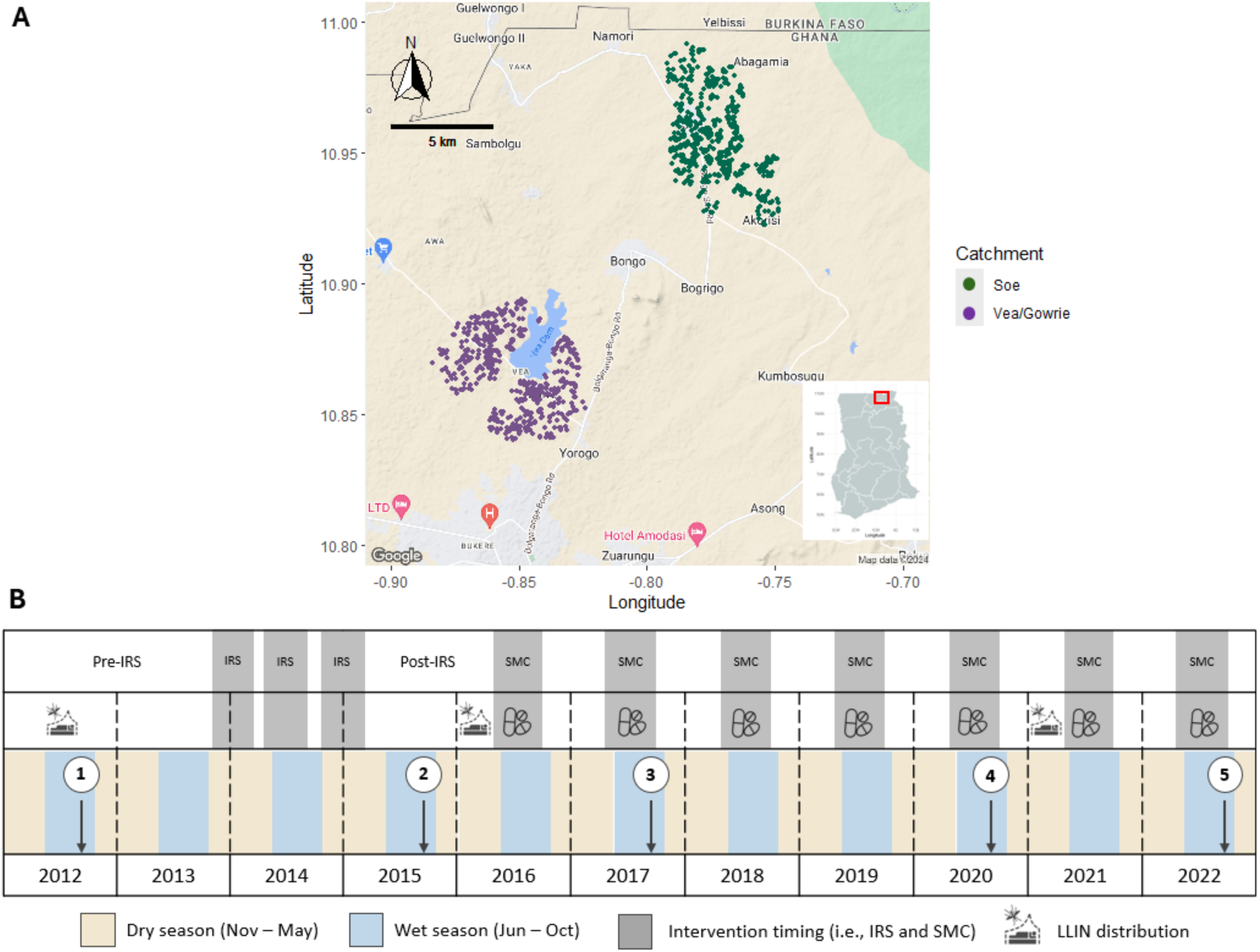
The study area and study design in Bongo District. (**A**) Study site locations in Bongo District, Ghana, showing the distribution of the compounds (i.e., households) included from the two catchment areas, Vea/Gowrie (purple, lower left) and Soe (green, upper right), which are ∼20-40 km apart. The location of Bongo District (red square) in the Upper East Region of northern Ghana is provided in the insert map (lower right). As the population in Bongo District resides in rural communities scattered throughout the district, only those compounds geolocated and included in this study are shown on the map. The map was drawn using the R packages *ggmap* [27], *malariaAtlas* [28], and *ggspatial* [29]. (**B**) Five age-stratified cross-sectional surveys (N = ∼2,000 participants per survey) were conducted in Bongo at the end of the wet seasons between 2012 and 2022 (white circles) in October 2012 (Survey 1, IRS), October 2015 (Survey 2, post-IRS), October 2017 (Survey 3, SMC), November 2020 (Survey 4, SMC), and October 2022 (Survey 5, SMC). Three rounds of IRS with non-pyrethroid insecticides were implemented between 2013-2015 as indicated. Starting in 2016, SMC with SPAQ, was administered to all children (< 5 years) during the wet season (i.e., high transmission). Both IRS and SMC were implemented against a background of widespread LLIN (impregnated with pyrethroid insecticides) usage which were distributed across Bongo District in 2012, 2016, and 2021 through universal coverage campaigns. For additional details on these interventions please see the Materials and Methods. Fig 1B was adapted from [12,30].

### Malaria control interventions

Between 2010 and 2012 and again in 2016 and 2021, LLINs impregnated with pyrethroid insecticides (i.e., PermaNet 2.0, Olyset, or DawaPlus 2.0) were mass distributed in Bongo District through universal coverage campaigns (UCC) by the National Malaria Elimination Programme (NMEP)/Ghana Health Service (GHS) [12,16–18,31]. For those years when no UCC were undertaken, LLINs were provided through routine services (e.g., antenatal clinics, school distributions, etc.) to maintain coverage [12,17].

Starting in 2013, three rounds of IRS with non-pyrethroid insecticides (i.e., organophosphate pirimiphos-methyl formulations) was implemented across the Upper East Region, including Bongo District, with the support of the AngloGold Ashanti Malaria Control Programme (AGAMal) and funding from the Global Fund [12,17,18,32]. The three rounds of IRS were implemented across Bongo District between 2013 and 2015: Round 1: October 2013 – January 2014, (Vectoguard 40WP), Round 2: May - July 2014 (Actellic 50EC), and finally Round 3: December 2014 - February 2015 (Actellic 300CS). Additional details on the IRS intervention have been previously described and published [12]

Following the discontinuation of this short-term IRS in 2015, SMC was rolled out across the Upper East Region starting in 2016 by Ghana’s NMEP with funding from the Global Fund [17]. SMC involves the monthly administration (i.e., ∼28-30 days apart) of a full treatment course of an antimalarial to all children between 3-59 months (i.e., < 5 years; SMC-eligible age group), regardless of infection status, for up to four months of the year during the wet or high-transmission season [33]. Like other countries of the Sahel sub-region of Africa where SMC was first deployed, the current drug of choice for SMC in Ghana is sulfadoxine-pyrimethamine plus amodiaquine (SPAQ)[34].

### Study design and sample collection

Using an interrupted time-series study design, five age-stratified cross-sectional surveys of ∼2,000 participating individuals per survey, were carried out between 2012 and 2022 to evaluate the impact of IRS and SMC in combination with LLINs on the prevalence and composition of the *Plasmodium* spp. reservoir, specifically the non-falciparum minor species *P. malariae*, *P. ovale* spp., and *P. vivax* (Fig 1B). These ∼2,000 participants of all ages (1-98 years) represent ∼15% of the total population that reside in the Vea/Gowrie and Soe catchment areas [19]. The five cross-sectional surveys were undertaken at the end of the wet seasons, when parasite prevalence would be expected to be highest and can be broken down into four phases (1) October 2012 (Survey 1) prior to IRS and SMC (i.e., baseline); (2) October 2015 (Survey 2) seven months after the final round of IRS; (3) October 2017 (Survey 3) 32 months after IRS was discontinued, but during the deployment of SMC to the younger children (< 5 years); and finally (4) November 2020 (Survey 4) and October 2022 (Survey 5) more than five years after IRS was withdrawn but following the continued deployment of SMC for five and seven years, respectively (Fig 1B).

During each survey, all participants completed a structured questionnaire to collect information on their demographics and socioeconomic characteristics, malaria prevention activities, recent clinical symptoms, blood pressure, body weight, axillary temperature, etc. To monitor for intervention use, information was collected from all participants on IRS coverage, LLIN usage the previous night, and antimalarial treatment in the previous two-weeks (i.e., participants that reported they were sick, sought treatment, and were provided with an antimalarial treatment) [12,19]. In addition, blood samples were collected for thick/thin blood films, rapid diagnostic tests (RDT), haemoglobin concentration, and as dried blood spots (DBS) on filter paper for laboratory-based molecular analyses. All participants who had a positive RDT and were febrile (temperature ≥ 37.5°C) were considered to be symptomatic for malaria and were provided with the appropriate care by clinical personnel at the local health centres via the Ministry of Health (MOH)/Ghana Health Service (GHS), following the current NMEP antimalarial treatment policy) [35,36]

This study enrolled 3,239 participants, across the five cross-sectional surveys, spanning all ages (median age: 12 years [IQR = 5-28; range = 1-98]) (Table 1). From these participants, 2,552 (78.8%; median age: 13 years [IQR = 7-32; range = 1-98]) were surveyed in more than one of the five cross-sectional surveys (range of surveys: 2 to 5) resulting in 9,006 DBS samples (92.9%) from the total 9,693 DBS samples collected and analysed using the laboratory-based molecular diagnostics. For the 687 (21.2%) participants that were only surveyed once in the five surveys considered here, 303 (44.1%) were from children newly enrolled in the final 2022 survey to maintain the age-stratified study design.

**Table 1.** Demographic characteristics of the study population during each study time point.

| Characteristics <sup>a</sup> | October 2012<br>(Survey 1, pre-IRS) | October 2015<br>(Survey 2, post-IRS) | October 2017<br>(Survey 3, SMC) | November 2020<br>(Survey 4, SMC) | October 2022<br>(Survey 5, SMC) |
| --- | --- | --- | --- | --- | --- |
| **Total number of participants** | 1,923 | 2,022 | 1,915 | 1,809 | 2,026 |
| **Age groups** |  |  |  |  |  |
| < 5 years | 271 (14.1) | 321 (15.9) | 295 (15.4) | 326 (18.0) | 393 (19.4) |
| 5-10 years | 480 (25.0) | 493 (24.4) | 417 (21.8) | 326 (18.0) | 445 (22.0) |
| 11-20 years | 413 (21.5) | 467 (23.1) | 489 (25.5) | 515 (28.5) | 513 (25.3) |
| ≥ 21years | 759 (39.5) | 741 (36.6) | 714 (37.3) | 642 (35.5) | 675 (33.3) |
| Age, years, median<br>(IQR; range) | 15<br>(7-36; 1-92) | 13<br>(7-35; 1-95) | 14<br>(8-35; 1-97) | 14<br>(7-35; 1-96) | 13<br>(6-31; 1-98) |
| **Sex** |  |  |  |  |  |
| Female | 1,031 (53.6) | 1,093 (54.1) | 1,036 (54.1) | 986 (54.5) | 1,119 (55.2) |
| Male | 892 (46.4) | 929 (45.9) | 879 (45.9) | 823 (45.5) | 907 (44.8) |
| **Catchment area** |  |  |  |  |  |
| Soe | 1,004 (52.2) | 1,022 (50.5) | 1,003 (52.4) | 935 (51.7) | 1,037 (51.2) |
| Vea/Gowrie | 919 (47.8) | 1,000 (49.5) | 912 (47.6) | 874 (48.3) | 989 (48.8) |
| **LLIN usage** (previous night) |  |  |  |  |  |
| Yes | 1,713 (89.1) | 1,831 (90.6) | 1,853 (96.8) | 1,690 (93.4) | 1,816 (89.6) |
| **Antimalarial treatment**<br>(previous two weeks) <sup>b</sup> |  |  |  |  |  |
| Treatment | 796 (41.4) | 298 (14.7) | 313 (16.3) | 346 (19.1) | 654 (32.3) |
| No treatment | 1,127 (58.6) | 1,645 (81.4) | 1,558 (81.4) | 1,437 (79.4) | 1,366 (67.4) |
| Don't know | 0 (0.0) | 79 (3.9) | 44 (2.3) | 26 (1.4) | 6 (0.3) |

Abbreviations: LLIN = long-lasting insecticidal net; IQR = Interquartile Range (25th–75th percentile); range = minimum–maximum values.
<sup>a</sup> Data reflects the number (% (n/N)) of participants surveyed.
<sup>b</sup> Indicates those participants who reported they were sick, sought treatment, and were provided with an antimalarial treatment in the previous two weeks. For those participants who didn't know if they were provided with an antimalarial treatment are coded as "Don't know".

### Molecular estimation of *Plasmodium* spp. prevalence

Genomic DNA (gDNA) was extracted from the DBS samples collected during each of the five surveys using the QIAamp DNA Mini Kit (QIAGEN, USA) as previously described [19]. All samples were then tested for *Plasmodium* spp. infections (i.e., *P. falciparum*, *P. malariae*, *P. ovale* spp., and *P. vivax*) using a nested PCR targeting the 18S ribosomal RNA gene (*18S rRNA*) [37–39] using previously primers and protocols with modifications [19]. The modified protocol and primer sequences for the species-specific *18S rRNA* PCR are available online on GitHub (https://github.com/UniMelb-Day-Lab/SpeciesSpecific_18S_rRNA_PCR). Using this laboratory-based molecular approach, we were able to identify all participants with single- or mixed-*Plasmodium* spp. infections (i.e., *P. falciparum*, *P. malariae*, *P. ovale* spp., and/or *P. vivax*). Since *P. ovale* was first identified by Stephens in 1922, it has been considered a single species. However, molecular studies have now revealed two genetically distinct forms, commonly referred to as *P. ovale curtisi* and *P. ovale wallikeri* [40]. Although the formal taxonomic classification of these species remains under debate [41–44] this study follows the prevailing convention in the literature by referring to them as *P. ovale curtisi* and *P. ovale wallikeri*, while collectively describing them as *P. ovale* spp.

### Statistical analysis

For the analyses, participants were categorized into defined age groups, sex, and catchment areas. Continuous variables are presented as medians with interquartile ranges (IQRs), and categorical variables are presented using prevalence with 95% confidence intervals (CIs). For these analyses, a single-species infection was defined as an infection with *P. falciparum, P. malariae, P. ovale* spp., or *P. vivax,* while a mixed-species infection was defined as an infection with *P. falciparum, P. malariae, P. ovale* spp., and/or *P. vivax.* Associations between the various interventions or study time points (i.e., 2012, 2015, 2017, 2020, and 2022) and the prevalence of each *Plasmodium* spp. (including single- and mixed-species infections) were estimated using multivariable logistic regression. Confounding variables identified a priori (age groups, sex, catchment area, LLIN usage, and antimalarial treatment in the previous two weeks) were adjusted for in the multivariable regression models. Effect modification by age group, identified a priori, was investigated using an interaction term between this variable and study time points (i.e., Surveys). Using this analysis, we encountered situations where there were no *Plasmodium* spp. infections (including single- and mixed-species). To address this issue, we employed Firth’s Bias-Reduced logistic regression. Since the majority of the study participants were re-assessed during each of the five cross-sectional surveys the cluster sandwich variance estimator was used to deal with the correlation of these repeated measurements. All statistical analyses were performed in R version 4.4.1 (2024-06-14) and RStudio version 2024.04.2 Build 764 [45] using base R, *tidyverse* [46], *epiR* [47], *stats* [45]*, gtsummary [48], nnet, clubSandwich* [49], and *logistf* [50].

## RESULTS

The study population characteristics for each of the five cross-sectional surveys or study time points (i.e., 2012, 2015, 2017, 2020, and 2022) are provided in Table 1. The study was designed to look at the asymptomatic reservoir in an age-stratified sample of the Bongo population in the two catchment areas (Vea/Gowrie and Soe). No significant differences between the proportion of participants surveyed in each of the catchment areas were observed. Analyses, unless otherwise stated, combined both catchment areas. Likewise, there was a similar proportion of female and male participants across the five surveys. As there was no difference in infection prevalence between the older age groups (i.e., 21-39 and ≥ 40 years), we combined these age groups and described them collectively as the adult population (≥ 21 years).

### Species-specific prevalence and species composition of infections

All DBS samples collected during each of the five study time points (N = 9,693) were processed, including gDNA extraction and species-specific *18S rRNA* PCR for *Plasmodium* spp. detection and identification. Among these samples analysed, 55.1% (N = 5,336) were positive for *P. falciparum*, 6.2% (N = 598) for *P. malariae*, and 2.8% (N = 267) for *P. ovale* spp., including both single- and mixed-*Plasmodium* spp. infections (Table 2). As expected, no samples were PCR-positive for *P. vivax*. Across all study time points between 2012 to 2022, single-*P. falciparum* infections were the most common, accounting for 76.2% to 95.4% of PCR-positive samples (Table 2). In contrast, single minor-species infections were rare, with *P. malariae* and *P. ovale* spp. each accounting for less than 2.1% and 0.8% of the PCR-positive samples, respectively (Table 2). Instead, most minor-species infections occurred as mixed infections with *P. falciparum*, the dominant species. Across the study time points mixed-*P. falciparum/P. malariae* infections ranged from 2.4% to 15.4%, while mixed-*P. falciparum/P. ovale* spp. infections ranged from 0.9% to 4.6% of the PCR-positive samples (Table 2). Triple-species infections, although observed, were uncommon throughout the study. Given the paucity of single minor-species infections, both single- and mixed-species infections were grouped together for *P. falciparum, P. malaria,* and *P. ovale* spp. in subsequent analyses (S1 Table).

**Table 2.** Number and percentage of single- and mixed-*Plasmodium* spp. infections as determined by the species-specific *18S rRNA* PCR during each study time point. See S1 Table for additional details.

|  | October 2012<br>(Survey 1, pre-IRS) | October 2015<sup>a</sup><br>(Survey 2, post-IRS) | October 2017<br>(Survey 3, SMC) | November 2020<br>(Survey 4, SMC) | October 2022<br>(Survey 5, SMC) |
| --- | --- | --- | --- | --- | --- |
| **Number of DBS tested** | 1,923 | 2,020 | 1,915 | 1,809 | 2,026 |
| **Total number of *Plasmodium* spp. infections<sup>b</sup>** | 1,420 (73.8) | 799 (39.6) | 1,227 (64.1) | 922 (51.0) | 1,050 (51.8) |
| ***Plasmodium* spp. single-species infections<sup>c</sup>** |  |  |  |  |  |
| *P. falciparum* | 1,082 (76.2) | 762 (95.4) | 1,101 (89.7) | 758 (82.2) | 910 (86.7) |
| *P. malariae* | 10 (0.7) | 10 (1.3) | 3 (0.2) | 19 (2.1) | 12 (1.1) |
| *P. ovale* spp. | 8 (0.6) | 1 (0.1) | 10 (0.8) | 6 (0.7) | 2 (0.2) |
| ***Plasmodium* spp. mixed-species infections<sup>c</sup>** |  |  |  |  |  |
| *P. falciparum*/*P. malariae* | 218 (15.4) | 19 (2.4) | 47 (3.8) | 98 (10.6) | 102 (9.7) |
| *P. falciparum*/*P. ovale* spp. | 66 (4.6) | 7 (0.9) | 61 (5.0) | 25 (2.7) | 21 (2.0) |
| *P. malariae*/*P. ovale* spp. | 0 (0.0) | 0 (0.0) | 0 (0.0) | 1 (0.1) | 0 (0.0) |
| *P. falciparum*/*P. malariae*/*P. ovale* spp. | 36 (2.5) | 0 (0.0) | 5 (0.4) | 15 (1.6) | 3 (0.3) |

Abbreviations: DBS = dried blood spot
<sup>a</sup> For the *Plasmodium* spp. infections in October 2015, there were 2,020 participants included in the analysis: participants were excluded when no DBS was available for PCR (N=2).
<sup>b</sup> Data reflect the total number (% (n/N)) of participants that were positive for *Plasmodium* spp. infections (single- and mixed-species infections) as determined by 18S rRNA PCR, relative to the total number of DBS tested.
<sup>c</sup> Data reflect the total number (% (n/N)) of participants that were positive for *Plasmodium* spp. infections (single- and mixed-species infections) as determined by 18S rRNA PCR, relative to the total number of *Plasmodium* spp. infections.

### Impact of the IRS and SMC interventions on *Plasmodium* spp. infections

When we investigated the impact of adding IRS with non-pyrethroid insecticides to LLINs impregnated with pyrethroid insecticides to reduce the size of the *Plasmodium* spp. reservoir, we found that the prevalence of infection in Bongo, including all species, declined from 73.8% at baseline in 2012 (pre-IRS) to 39.6% in 2015 (Table 2, S1 Table) following the final round of IRS, which decreased transmission intensity of *P. falciparum* by > 90%, as measured by the entomological inoculation rate [12]. Overall, IRS reduced the prevalence of all *Plasmodium* spp., while *P. falciparum* remained the major species present (Fig 2A, Table 2, S1 Table). This decline in prevalence was short lived and in 2017, 32 months after IRS was discontinued, but during the deployment of SMC to all children < 5 years, *Plasmodium* spp. prevalence rebounded to 64.1% (Table 2). Finally in 2020 and 2022, five and seven years after IRS was withdrawn, but with sustained deployment of SMC, *Plasmodium* spp. prevalence, although lower than the 2017 rebound, remained high, with 51.0% and 51.8% of the population infected in 2020 and 2022, respectively (Table 2). By monitoring this parasite reservoir longitudinally, we found that although the prevalence of *P. falciparum* rebounded following the discontinuation of IRS, the minor-species infections, *P. malariae* and *P. ovale* spp., did not follow the same pattern as that observed for *P. falciparum*, both in the population (Fig 2A, S1 Table) and for each age group (Fig 4A, S1 Table).

**Fig 2.**
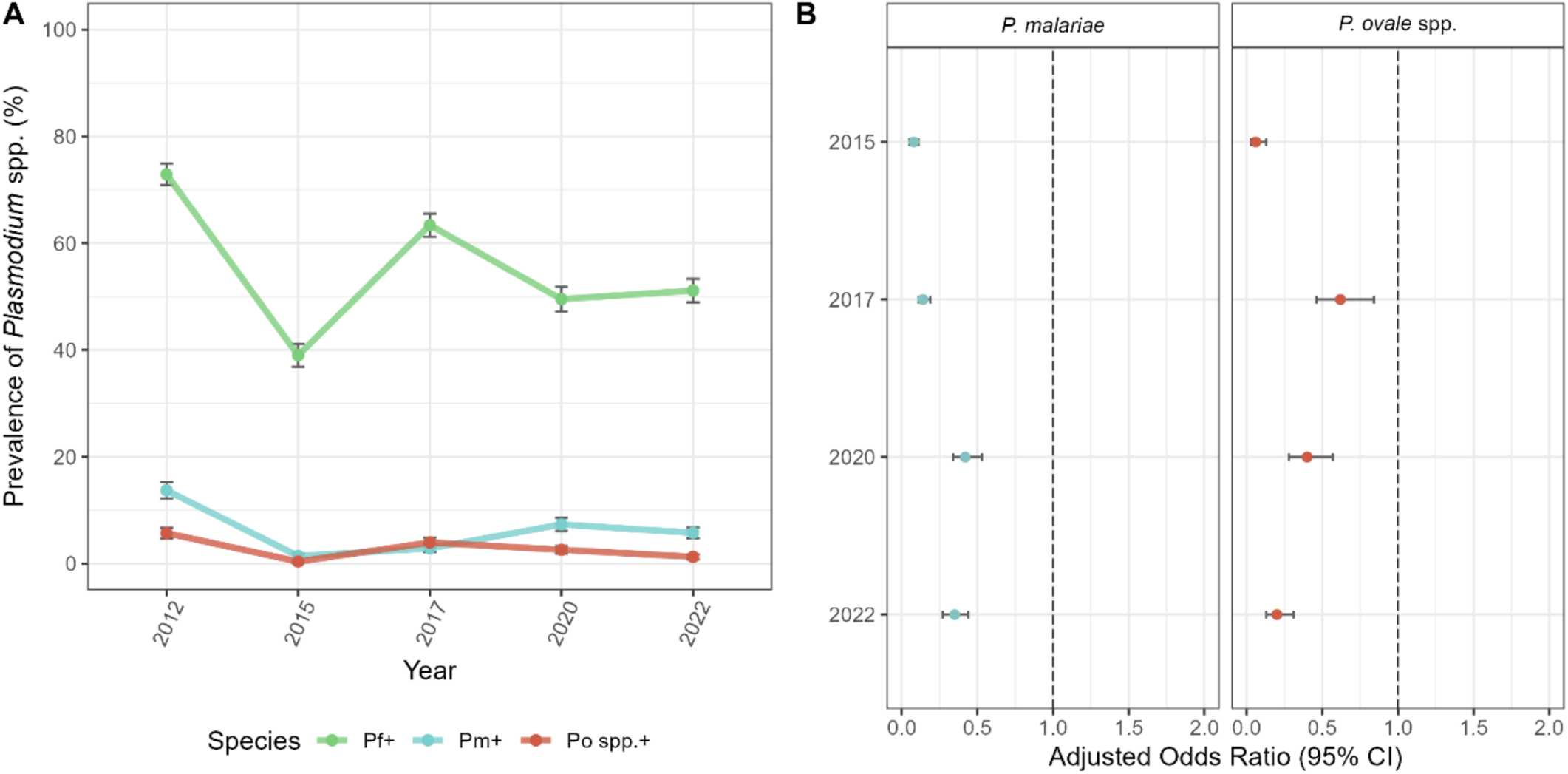
Changes in *Plasmodium* spp. prevalence between 2012 and 2022, following the implementation of IRS and SMC. (**A**) Prevalence of *P. falciparum* (green)*, P. malariae* (blue), and *P. ovale* spp. (red) infections (including single- and mixed-species infections) during each study time point (S1 Table). (**B**) Forest plots of the adjusted odds ratios for the association between the study time points for *P. malariae* (blue, left panel) and *P. ovale* spp. (red, right panel) infection prevalence compared to 2012 (i.e., baseline pre-IRS reference). Error bars (vertical: prevalence and horizontal: aOR) in the panels represent the upper and lower limits of the 95% confidence intervals (95% CI). For additional details see S2 Table.

### Impact of the IRS and SMC on minor-*Plasmodium* spp. infections

Prior to implementation of the IRS and SMC interventions, Table 2 shows *P. falciparum* as the dominant species with a substantial hidden reservoir of both *P. malariae* and *P. ovale* spp. with an overall prevalence of 13.7% and 5.7%, respectively, in single- and mixed-species infections in all ages (Table 2, S1 Table). Following the addition of IRS to LLINs, prevalence dropped for both *P. malariae* (13.7% to 1.4%, respectively) and *P. ovale* spp. (5.7% to 0.4%, respectively) between 2012 and 2015 (Fig 2A, S1 Table). In fact, in 2015, seven months after the final round of IRS, participants were 92% (aOR = 0.08 [95% CI: 0.05-0.11], p-value < 0.001) and 94% (aOR = 0.06 [95% CI: 0.03-0.13], p-value < 0.001) less likely to have a *P. malariae* and *P. ovale* spp. infection, respectively, compared to the 2012 baseline (Fig 2B, S2 Table).

Despite this decline, in 2017, 32 months after the discontinuation of IRS, but during SMC, the prevalence of *P. malariae* (2.9%) and *P. ovale* spp. (4.0%) rebounded (Fig 2A, S1 Table). In the adjusted model, the odds of infection for the minor species increased by ∼2-fold for *P. malariae* (aOR = 0.14 [95% CI: 0.11-0.19], p-value < 0.001) and by ∼10-fold for *P. ovale* spp. (aOR = 0.62 [95% CI: 0.46-0.84] p-value = 0.002), compared to 2015 (Fig 2B, S2 Table). It is important to point out that although prevalence rebounded in 2017 for the minor species, prevalence did not fully recover, with the population still being significantly less likely to have a *P. malariae* (aOR = 0.14 [95% CI: 0.11-0.19], p-value < 0.001) or *P. ovale* spp. (aOR = 0.62 [95% CI: 0.46-0.84], p-value = 0.002) infection, compared to the 2012 baseline (Fig 2B, S2 Table). In this study we found that although IRS significantly impacted the minor species, they did not rebound at the same rate once IRS was withdrawn. In fact, the prevalence of *P. malariae* did not recover to the same extent compared to *P. ovale* spp., in 2017 (Fig 2A, S1 and S3 Tables) possibly due to the ability of *P. ovale* spp. to relapse.

When we investigated the Bongo population (i.e., in all ages) in 2020, five years post-IRS, but during the continued deployment of SMC, the prevalence of *P. malariae* continued to recover (7.4%), while the prevalence of *P. ovale* spp. actually declined (2.6%) after rebounding in 2017 (Fig 2A, S1 Table). According to the adjusted models, although the odds of a *P. malariae* and *P. ovale* spp. infection in 2020 were still significantly lower than 2012, the odds of a *P. malariae* infection (aOR = 0.42 [95% CI: 0.34-0.53] p-value < 0.001) continued to increase over time compared to 2017, while for *P. ovale* spp., it declined (aOR = 0.40 [95% CI: 0.28-0.57], p-value < 0.001) (Fig 2B, S2 Table).

Finally, when we looked at the population in 2022, more than seven years after IRS was discontinued, but with sustained SMC, both *P. malariae* prevalence (5.8%) and odds of infection (aOR = 0.35 [95% CI: 0.27-0.44], p-value < 0.001) decreased slightly compared to baseline in 2012 (Fig 2B, S2 Table). For *P. ovale* spp., prevalence continued to decline (1.3%) in 2022 along with the odds of infection (aOR = 0.20 [95% CI: 0.13-0.31], p-value < 0.001) (Fig 2B, S2 Table). Overall, although the prevalence of the minor-species infections rebounded in the population following the discontinuation of IRS, albeit delayed for *P. malariae*, neither minor species fully recovered relative to the 2012 baseline (i.e., prior to the sequential IRS and SMC interventions).

### Age-specific changes in the minor-*Plasmodium* spp. infections following IRS and SMC

This study was designed to evaluate the impact of IRS and SMC interventions on the prevalence of *Plasmodium* spp. infection by age. This is particularly important given that SMC is a chemoprevention strategy targeted at children < 5 years starting in 2016, once IRS was discontinued. For all species, prevalence of infection during each study time point remained highest in the older children (5-10 years) and adolescents (11-20 years) (Fig 3A, S1 Table). In 2015, following IRS, prevalence declined, with participants in each age group being ≥ 76% and ≥ 89% less likely to have a *P. malariae* and *P. ovale* spp. infection, respectively, compared to 2012 (p-values <0.001) (Figs 4B and 4C, S3 and S4 Tables). Although the prevalence of the minor species infections declined in all age groups following IRS, the greatest reductions were observed among the younger (< 5 years) and older (5-10 years) children surveyed. In fact, in 2015, seven months after the final round of IRS, younger and older children were ≥ 97% and 96% less likely to have a *P. malariae* and *P. ovale* spp. infection, respectively, compared to the 2012 baseline (p-values < 0.001) (Figs 4B and 4C, S3 and S4 Tables).

**Fig 3.**
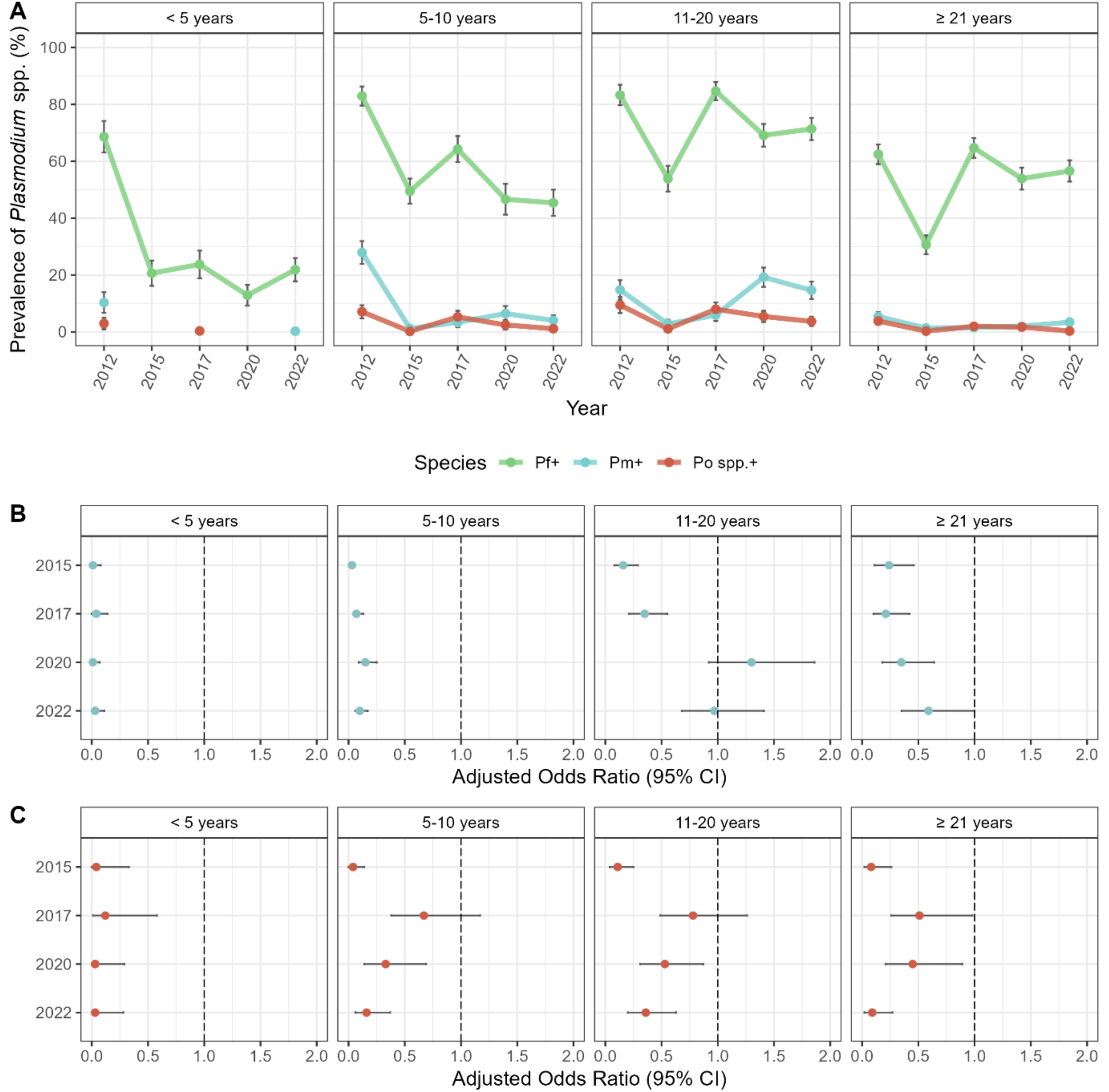
Age specific changes in *Plasmodium* spp. prevalence between 2012 and 2022, following the implementation of IRS and SMC. (**A**) Prevalence of *P. falciparum* (green)*, P. malariae* (blue), and *P. ovale* spp. (red) infections (including single- and mixed-species infections) by age group during each study time point (S1 Table). Forest plots representing the adjusted odds ratios for the associations between the study time points for (**B**) *P. malariae* (blue) and (**C**) *P. ovale* spp. (red) infection prevalence in each age group (years) compared to 2012 (i.e., baseline pre-IRS reference). Error bars (vertical: prevalence and horizontal: aOR) in the panels represent the upper and lower limits of the 95% confidence intervals (95% CI). For additional details see S3 and S4 Tables.

In 2017, 32 months after the discontinuation of IRS, minor species prevalence in the younger children (< 5 years) targeted by SMC, remained significantly lower for both *P. malariae* (aOR = 0.04 [95% CI: 0.00 – 0.14], p-value < 0.001) and *P. ovale* spp. (aOR = 0.12 [95% CI: 0.01 – 0.58], p-value = 0.007) compared to 2012 (Figs 4B and 4C, S3 and S4 Tables). Meanwhile for the older children (5-10 years), adolescents (11-20 years), and adults (≥ 21 years) that were all ineligible for SMC, the prevalence of minor-species infections rebounded, although once again *P. malariae* did not recover as quickly compared to *P. ovale* spp. (Fig 3A, S1 Table). In fact, further analysis of effect modification by age showed that while the odds of a *P. ovale* spp. infection for the older age groups (i.e., 5-10, 11-20, and ≥ 21 years) rebounded and was comparable to that observed in 2012 (Fig 3C, S4 Table), this was not the case for *P. malariae* (Fig 3B, S3 Table). In fact, the odds of a *P. malariae* infection for the older age groups remained significantly lower in 2017 compared to 2012, despite the discontinuation of IRS (Fig 3B, S3 Table).

During 2020 and 2022, the minor species were rarely detected in the younger children (< 5 years) eligible for SMC, with no such infections identified in 2020 and only one *P. malariae* infection detected in 2022 (Fig 3A, S1 Table). In both study time points, the majority of *P. malariae* and *P. ovale* spp. infections were found in older children (5-10 years) and adolescents (11-20 years) (Fig 3A, S1 Table). Although the prevalence of *P. ovale* spp. declined for the older age groups (i.e., 5-10, 11-20, and ≥ 21 years) starting in 2020, the prevalence of *P. malariae* increased (Fig 3A, S1 Table). When we examined this trend in more detail, we found that this delayed recovery of *P. malariae* was most noticeable for the adolescents (11-20 years) (Fig 3A, S1 Table) where the odds of infection in 2020 (aOR = 1.30 [95% CI: 0.92-1.86], p-value = 0.14) was comparable to baseline in 2012 (Fig 3B, S3 Table). In fact, by 2022, *P. malariae* prevalence had fully recovered for the adolescent (11-20 years) and adult (≥ 21 years) age groups (Figs 4A and 4B, S2 and S3 Tables). This however was not the case of the younger (< 5 years) and older children (5-10 years). In contrast to this delayed rebound for *P. malariae,* the odds of *P. ovale* spp. infections in 2022 continued to decline across all age groups from the rebound observed in 2017 (Fig 3C, S4 Table).

### Impact of the IRS and SMC on single- and mixed-*Plasmodium* spp. infections

Data on the proportions of single- and mixed-species infections during each study time point were further analysed to see how these changes following the sequential IRS and SMC interventions. Even after these interventions were deployed, *P. falciparum* remained the dominant species in Bongo. Across the five study time points between 2012 and 2022, the *P. falciparum* proportion, as a single-species infection, was greater than 0.75 (Fig S1A, Table 2). On the other hand, mixed-*P. falciparum* infections declined in 2015, seven months after the final round of IRS, compared to the baseline in 2012 (Fig S1A, Table 2). From 2017 to 2022, as transmission intensity recovered following the discontinuation of IRS, the proportion of mixed *P. falciparum* infections steadily increased from 0.03 in 2015 to 0.12 in 2022 (Fig S1A, Table 2).

When we examined the proportion of single- vs. mixed-species infection patterns based on age, we observed that mixed *P. malariae* and *P. ovale* spp. infections occurred mostly in the older children (5-10 years) and adolescents (11–20 years) during each study time point including the 2012 baseline (i.e., prior to the sequential IRS and SMC interventions) (Figs S1D, S1E, and S1F). In contrast, the single-*P. malariae* and *P. ovale* spp. infections were found mainly in the adults (≥ 21 years) (Figs S1E and S1F). This is consistent with the hypothesis that as immunity to *P. falciparum* increases, the prevalence of minor species increases such that the peak prevalence of the minor species and dominant species do not coincide [51].

This pattern holds even with intervention. In particular, we draw attention to increases in single-species infections with *P. malariae* and *P. ovale* spp., especially after IRS in adolescents (11-20 years) and adults (≥ 21 years) and to a lesser extent to older children (5-10 years) (Figs S1E and S1F). A notable increase in proportion of *P. malariae* occurs in 2015 when the prevalence of *P. falciparum* is the lowest.

## DISCUSSION

This study is the first longitudinal study to assess the impact of two sequential interventions, IRS and SMC in combination with widespread LLIN usage on the minor *Plasmodium* spp. in the reservoir of asymptomatic infection in Ghana and indeed West Africa. Employing species-specific molecular diagnostics, we observed a high prevalence of submicroscopic *P. malariae* at 13.7% and *P. ovale spp*. of 5.4% occurring largely as mixed infections with *P. falciparum* at baseline in 2012. The peak prevalences of these minor species occurred in older children (5-10 years) and adolescents (11-20 years) with detectable infections in both younger children (< 5 years) and adults (≥ 21 years). Given the implementation of control interventions across districts in northern Ghana, although often variable as influenced by shifts in funding and NMEP priorities, such high prevalences of submicroscopic minor *Plasmodium* spp. infections were more common than we might have expected.

From the baseline in 2012 (pre-IRS) we were able to assess the efficacy of sequential IRS and SMC interventions. Longitudinal data shows, without doubt, the effectiveness of the short-term IRS against all *Plasmodium* spp., even reducing the prevalence of minor *Plasmodium* spp. to near zero. Upon discontinuation of IRS, age-specific analyses showed a rebound in the prevalence of *P. falciparum* in 2017 (i.e., 32 months post-IRS), to levels higher than the 2012 baseline in adolescents and adults [20]. A similar age-specific shift, characterized by higher prevalence of minor *Plasmodium* spp. infections in adolescents and adults, was observed in 2020 after the initial rebound in 2017 following the withdrawal of IRS and five consecutive years of SMC. These age-shifts in prevalence of all *Plasmodium* spp. are likely due to loss of acquired immunity in all ages during the short-term IRS and SMC targeting children < 5 years. The older children showed a similar pattern, although with a relatively smaller increase in prevalence for both *P. falciparum* and minor *Plasmodium* spp. This can be explained by children who aged out of the < 5 years age group during the study; having received SMC previously, they may have reduced immunity and thus be more susceptible to infection. Additionally, these data could indicate “leakage” of SMC to older children outside the SMC-eligible age range, suggesting administration to children not formally targeted by the NMEP [52]. These upward age-specific shifts in prevalence make clear the importance of molecular surveillance to detect low-density asymptomatic infections in all ages. This is not the typical practice in Ghana nor other countries implementing these interventions in West Africa. Instead, the common practice is to undertake surveillance by focusing on children aged 2-10 years, as seen in the Malaria Atlas Project [1].

The reduction in the prevalence of all the *Plasmodium* spp. in the younger children proves that SMC, in combination with LLINs, is effective at protecting this target age group. These results indicate that the benefits of SMC could be expanded by increasing the ages that are eligible, specifically older children and adolescents. They have been shown in this study and others to be vulnerable and act as reservoirs for asymptomatic infections not only *P. falciparum* but also minor *Plasmodium* spp. [5,22,23]. Moreover, non-falciparum infections are mainly found at low-densities and are asymptomatic [53]. By expanding the SMC-eligible age range, the reservoir of *Plasmodium* spp. would be better controlled with targeted chemoprevention.

WHO recommends the combination SPAQ for SMC in areas with highly seasonal *P. falciparum* transmission, as it provides protection for ∼28 days, is well tolerated, relatively inexpensive, and has demonstrated appropriate efficacy in Africa [54]. Recently, there have been reports of moderate to high prevalence of mutations in the *P. ovale* spp. dihydrofolate reductase gene (*dhfr)*, associated with resistance to pyrimethamine, in isolates from Central and East Africa [55], as well as travellers returning from Africa [56] In this study, although we did not test for antimalarial resistance markers in minor *Plasmodium* spp., we found no evidence of emerging resistance to SPAQ after seven consecutive years of SMC implementation. This is supported by the sustained reduction in infection prevalence within the SMC target age group. Among the 1,014 SMC-eligible children (< 5 years) examined between 2017 and 2022, up to seven years after the introduction of SMC, only two *P. malariae* infections (2017 and 2022) and one *P. ovale* spp. (2017) infections were observed. Although we did not observe emergence of *P. malariae* in this age group, a need for monitoring drug resistance mutations in the non-falciparum species in this area remains.

Looking at age-specific changes in the proportions of each species (Fig S1), complex changes are seen in response to the sequential IRS and SMC interventions. A clear pattern to emerge is that the proportion of single-*P. malariae* and *P. ovale* spp. infections increases after the IRS intervention (2015), especially among the adolescents and adults, and also with age after the withdrawal of IRS (i.e., from 2017 onwards). This speaks to a potential interaction with the dominant species *P. falciparum* where *P. falciparum* density decreases with age and also after the IRS intervention [12,19]. If there is a carrying capacity for total *Plasmodium* spp. parasite numbers, as proposed by Bruce et al. [51], a decline in *P. falciparum* numbers would allow for an increase in density by minor *Plasmodium* spp.

The highly sensitive species-specific PCR targeting the *18S rRNA* gene [37–39] proved suitable to identify submicroscopic infections from DBS samples. The utility of quantitative real-time PCR for molecular epidemiological surveillance of these minor-species infections is questionable for National Malaria Programmes operating in moderate- to high-transmission settings, given the increased cost (e.g., specialized equipment, reagents, etc.). More detailed metagenomic analyses of variable blood volumes has shown that analysing gDNA from DBS samples is underestimating the prevalence of minor-species infections by a factor of 1.93× for *P. malariae* and 2.50× for *P. ovale* spp. [57]. We did not discriminate between the non-recombining sympatric species *P. ovale curtisi* and *P. ovale wallikeri*, recognised as distinct since 2010 [40]. This information will be reported separately.

In summary, we have uncovered a substantial “hidden reservoir” of submicroscopic *P. malariae* and *P. ovale* spp. infections across all ages in Bongo District in the Upper East Region of northern Ghana. These infections reduced in prevalence in response to the sequential IRS and SMC interventions implemented to target *P. falciparum*. From our data we make the case that the reservoir of infection should be monitored by molecular surveillance of *P. falciparum* and minor-*Plasmodium* spp. as high-burden SSA countries, like Ghana, advance towards malaria elimination.

## ACKNOWLEDGEMENTS

We wish to thank the participants, communities, traditional leadership, and the Ghana Health Service in Bongo District, Ghana for their willingness to participate in this study. We would like to thank the field teams in Bongo for their technical assistance in the field, as well as the laboratory personnel at the Navrongo Health Research Centre for their expertise and for undertaking the sample collections and parasitological assessments. We thank Mercedes Pascual (Department of Biology and Department of Environmental Studies, New York University, New York, US) for valuable input on this work and for her role as a multi-principal investigator in securing the funding that supported this research (see Funding for additional details).

## DATA AVAILABILITY STATEMENT

All data associated with this study are available in the manuscript, supplement, or upon reasonable request by contacting the Malaria Reservoir Study Team, represented by the corresponding author, Prof. Karen Day, to discuss how these data will be utilized for academic or research purposes and, if appropriate, to explore opportunities for collaboration.

## BENEFIT-SHARING STATEMENT

A research collaboration was developed with scientists from Ghana based at the Navrongo Health Research Centre and the Noguchi Memorial Institute for Medical Research. All collaborators are included as co-authors and the relevant results from the research has been shared with participants, key stakeholders, traditional leadership, and the local community (i.e., Paramount Chief of Bongo District, divisional Chiefs, Queen Mothers, and community members), the Bongo District and Upper East Regional Health Directorates, as well as Ghana National Malaria Elimination Programme. Before this research was undertaken, informed consent was sought and obtained from the key stakeholders, traditional leadership, and the local community in Bongo District. In addition, members of the local community were trained as field workers and were directly involved in liaising with the community and in the collection of the study data. The contribution of these individuals to this research is described in the Acknowledgements. This research addresses a priority concern regarding malaria control and the impact of interventions. These concerns are relevant for both the local community in Bongo District, as well as for the National Malaria Elimination Programme in Ghana.

## AUTHOR CONTRIBUTIONS

**Conceptualization:** KPD

**Data Curation:** KET, OB, DCA

**Formal Analysis:** CART, KET

**Funding Acquisition:** KAK, ARO, KPD

**Investigation:** CART, KET, OB, SLD, DCA

**Project Administration:** KET, OB, ARO, POA, KPD

**Resources:** ARO, POA, KPD

**Supervision:** KET, OB, ARO, POA, KPD

**Validation:** CART, KET

**Visualization:** CART, KET

**Writing – Original Draft Preparation:** CART, KET, KPD

**Writing – Review & Editing:** OB, SLD, DCA, KAK, ARO, POA

## SUPPORTING INFORMATION CAPTIONS

**S1 Table.** Parasitological characteristics of the *Plasmodium* spp. infections (including single- and mixed-species infections) during each study time point.

| Parasitological parameters | October 2012<br>(Survey 1, pre-IRS) | October 2015<sup>a</sup><br>(Survey 2, post-IRS) | October 2017<br>(Survey 3, SMC) | November 2020<br>(Survey 4, SMC) | October 2022<br>(Survey 5, SMC) |
| --- | --- | --- | --- | --- | --- |
| **Number of DBS tested** | 1,923 | 2,020 | 1,915 | 1,809 | 2,026 |
| ***P. falciparum* prevalence** (including single- and mixed-species infections) |  |  |  |  |  |
| **All<sup>b</sup>** | **1,402 (72.9)** | **788 (39.0)** | **1,214 (63.4)** | **896 (49.5)** | **1,036 (51.1)** |
| **Age groups<sup>c</sup>** |  |  |  |  |  |
| < 5 years | 186 (68.6) | 66 (20.6) | 70 (23.7) | 42 (12.9) | 86 (21.9) |
| 5-10 years | 398 (82.9) | 244 (49.5) | 268 (64.3) | 152 (46.6) | 202 (45.4) |
| 11-20 years | 344 (83.3) | 251 (53.9) | 414 (84.7) | 356 (69.1) | 366 (71.3) |
| ≥ 21 years | 474 (62.5) | 227 (30.6) | 462 (64.7) | 346 (53.9) | 382 (56.6) |
| **Sex<sup>c</sup>** |  |  |  |  |  |
| Female | 711 (69.0) | 389 (35.6) | 646 (62.4) | 490 (49.7) | 543 (48.5) |
| Male | 691 (77.5) | 399 (43.0) | 568 (64.6) | 406 (49.3) | 493 (54.4) |
| **Catchment area<sup>c</sup>** |  |  |  |  |  |
| Soe | 801 (79.8) | 440 (43.1) | 685 (68.3) | 478 (51.1) | 528 (50.9) |
| Vea/Gowrie | 601 (65.4) | 348 (34.8) | 529 (58.0) | 418 (47.8) | 508 (51.4) |
| ***P. malariae* prevalence** (including single- and mixed-species infections) |  |  |  |  |  |
| **All<sup>b</sup>** | **264 (13.7)** | **29 (1.4)** | **55 (2.9)** | **133 (7.4)** | **117 (5.8)** |
| **Age groups<sup>c</sup>** |  |  |  |  |  |
| < 5 years | 28 (10.3) | 0 (0.0) | 1 (0.3) | 0 (0.0) | 1 (0.3) |
| 6-10 years | 134 (27.9) | 6 (1.2) | 14 (3.4) | 21 (6.4) | 18 (4.0) |
| 11-20 years | 61 (14.8) | 13 (2.8) | 29 (5.9) | 99 (19.2) | 75 (14.6) |
| ≥ 21 years | 41 (5.4) | 10 (1.3) | 11 (1.5) | 13 (2.0) | 23 (3.4) |
| **Sex<sup>c</sup>** |  |  |  |  |  |
| Female | 123 (11.9) | 13 (1.2) | 25 (2.4) | 56 (5.7) | 42 (3.8) |
| Male | 141 (15.8) | 16 (1.7) | 30 (3.4) | 77 (9.4) | 75 (8.3) |
| **Catchment area<sup>c</sup>** |  |  |  |  |  |
| Soe | 166 (16.5) | 19 (1.9) | 42 (4.2) | 70 (7.5) | 50 (4.8) |
| Vea/Gowrie | 98 (10.7) | 10 (1.0) | 13 (1.4) | 63 (7.2) | 67 (6.8) |
| ***P. ovale* spp. prevalence** (including single- and mixed-species infections) |  |  |  |  |  |
| **All<sup>b</sup>** | **110 (5.7)** | **8 (0.4)** | **76 (4.0)** | **47 (2.6)** | **26 (1.3)** |
| **Age groups<sup>c</sup>** |  |  |  |  |  |
| < 5 years | 8 (3.0) | 0 (0.0) | 1 (0.3) | 0 (0.0) | 0 (0.0) |
| 6-10 years | 34 (7.1) | 1 (0.2) | 22 (5.3) | 8 (2.5) | 5 (1.1) |
| 11-20 years | 39 (9.4) | 5 (1.1) | 39 (8.0) | 28 (5.4) | 19 (3.7) |
| ≥ 21 years | 29 (3.8) | 2 (0.3) | 14 (2.0) | 11 (1.7) | 2 (0.3) |
| **Sex<sup>c</sup>** |  |  |  |  |  |
| Female | 52 (5.0) | 3 (0.3) | 32 (3.1) | 18 (1.8) | 11 (1.0) |
| Male | 58 (6.5) | 5 (0.5) | 44 (5.0) | 29 (3.5) | 15 (1.7) |
| **Catchment area<sup>c</sup>** |  |  |  |  |  |
| Soe | 66 (6.6) | 7 (0.7) | 57 (5.7) | 24 (2.6) | 15 (1.4) |
| Vea/Gowrie | 44 (4.8) | 1 (0.1) | 19 (2.1) | 23 (2.6) | 11 (1.1) |

<sup>a</sup> For the *Plasmodium* spp. infections in October 2015, there were 2,020 participants included in the analysis: participants were excluded when no DBS was available for PCR (N=2).
<sup>b</sup> Data reflects the number (% (n/N)) of participants sampled that were positive for *P. falciparum*, *P. malariae*, or *P. ovale* spp. (including single- and mixed-species infections) as determined by *18S rRNA* PCR, relative to the total number of DBS samples tested. See Table 2 for the breakdown of the *Plasmodium* spp. groupings for single and mixed infections.
<sup>c</sup> Data reflects the number (% (n/N)) of participants sampled that were positive for *P. falciparum*, *P. malariae*, or *P. ovale* spp. (including single- and mixed-species infections) as determined by *18S rRNA* PCR, relative to the total number of DBS samples tested for each age group, sex, and catchment area. See Table 1 for the demographic breakdown by age group, sex, and catchment area.

**S2 Table.** Association between the study time points and *P. malariae* and *P. ovale* spp. infection prevalence using the species-specific *18S rRNA* PCR.

| Factor | *P. malariae* infection<br>(including single- and mixed-species infection) <sup>a</sup> |  |  |  | *P. ovale* spp. infection<br>(including single- and mixed-species infection) <sup>a</sup> |  |  |  |
| --- | --- | --- | --- | --- | --- | --- | --- | --- |
|  | Unadjusted<br>OR (95% CI) | *p*-value | Adjusted <sup>b</sup><br>aOR (95% CI) | *p*-value | Unadjusted<br>OR (95% CI) | *p*-value | Adjusted <sup>b</sup><br>aOR (95% CI) | *p*-value |
| **Study time point (Survey, Intervention)** |  |  |  |  |  |  |  |  |
| October 2012 (Survey 1, pre-IRS) | 1.00 | - | 1.00 | - | 1.00 | - | 1.00 | - |
| October 2015 (Survey 2, post-IRS) | 0.10 (0.07-0.14) | **<0.001** | 0.08 (0.05-0.11) | **<0.001** | 0.07 (0.03-0.14) | **<0.001** | 0.06 (0.03-0.13) | **<0.001** |
| October 2017 (Survey 3, SMC) | 0.18 (0.14-0.24) | **<0.001** | 0.14 (0.11-0.19) | **<0.001** | 0.70 (0.52-0.94) | **0.018** | 0.62 (0.46-0.84) | **0.002** |
| November 2020 (Survey 4, SMC) | 0.50 (0.41-0.62) | **<0.001** | 0.42 (0.34-0.53) | **<0.001** | 0.45 (0.31-0.63) | **<0.001** | 0.40 (0.28-0.57) | **<0.001** |
| October 2022 (Survey 5, SMC) | 0.39 (0.31-0.48) | **<0.001** | 0.35 (0.27-0.44) | **<0.001** | 0.21 (0.14-0.33) | **<0.001** | 0.20 (0.13-0.31) | **<0.001** |
| **Age groups** |  |  |  |  |  |  |  |  |
| < 5 years | 1.00 | - | 1.00 | - | 1.00 | - | 1.00 | - |
| 5-10 years | 5.13 (3.47-7.59) | **<0.001** | 5.14 (3.50-7.57) | **<0.001** | 5.92 (2.94-11.9) | **<0.001** | 5.57 (2.74-11.3) | **<0.001** |
| 11-20 years | 6.77 (4.66-9.84) | **<0.001** | 6.49 (4.46-9.44) | **<0.001** | 10.1 (5.09-19.9) | **<0.001** | 9.66 (4.84-19.3) | **<0.001** |
| ≥ 21 years | 1.48 (0.97-2.25) | 0.067 | 1.36 (0.89-2.08) | 0.2 | 2.99 (1.48-6.04) | **0.002** | 2.81 (1.37-5.77) | **0.005** |
| **Sex** |  |  |  |  |  |  |  |  |
| Female | 1.00 | - | 1.00 | - | 1.00 | - | 1.00 | - |
| Male | 1.60 (1.33-1.92) | **<0.001** | 1.39 (1.15-1.68) | **<0.001** | 1.56 (1.22-1.99) | **<0.001** | 1.41 (1.10-1.81) | **0.007** |
| **Catchment area** |  |  |  |  |  |  |  |  |
| Soe | 1.00 | - | 1.00 | - | 1.00 | - | 1.00 | - |
| Vea/Gowrie | 0.75 (0.62-0.90) | **0.002** | 0.70 (0.58-0.85) | **<0.001** | 0.60 (0.46-0.77) | **<0.001** | 0.57 (0.44-0.74) | **<0.001** |
| **LLIN usage (previous night)** |  |  |  |  |  |  |  |  |
| No | 1.00 | - | 1.00 | - | 1.00 | - | 1.00 | - |
| Yes | 0.71 (0.54-0.92) | **0.011** | 1.02 (0.77-1.36) | 0.9 | 0.86 (0.56-1.30) | 0.5 | 1.08 (0.70-1.68) | 0.7 |
| **Antimalarial treatment (previous two weeks) <sup>c</sup>** |  |  |  |  |  |  |  |  |
| No treatment | 1.00 | - | 1.00 | - | 1.00 | - | 1.00 | - |
| Treatment | 0.77 (0.63-0.94) | **0.011** | 0.55 (0.44-0.68) | **<0.001** | 0.84 (0.62-1.12) | 0.2 | 0.76 (0.56-1.03) | 0.076 |

OR = odds ratio; aOR = adjusted odds ratio; CI = confidence interval; LLIN = long-lasting insecticidal net
<sup>a</sup> Participants that were sick, sought treatment, but didn't know if they were provided with an antimalarial treatment in the previous two weeks were excluded from the model: October 2015 (N=79; 3.9%); October 2017 (N=44; 2.3%); November 2020 (N=26; 1.4%); and October 2022 (N=6; 0.3%).
<sup>b</sup> Age groups, sex, catchment area, LLIN usage (previous night), and antimalarial treatment (previous two weeks) are adjusted for in the multivariable logistic regression model.
<sup>c</sup> Indicates those participants who reported they were sick, sought treatment, and were provided with an antimalarial treatment in the previous two weeks. For those participants who didn't know if they were provided with an antimalarial treatment are coded as "Don't know".

**S3 Table.**
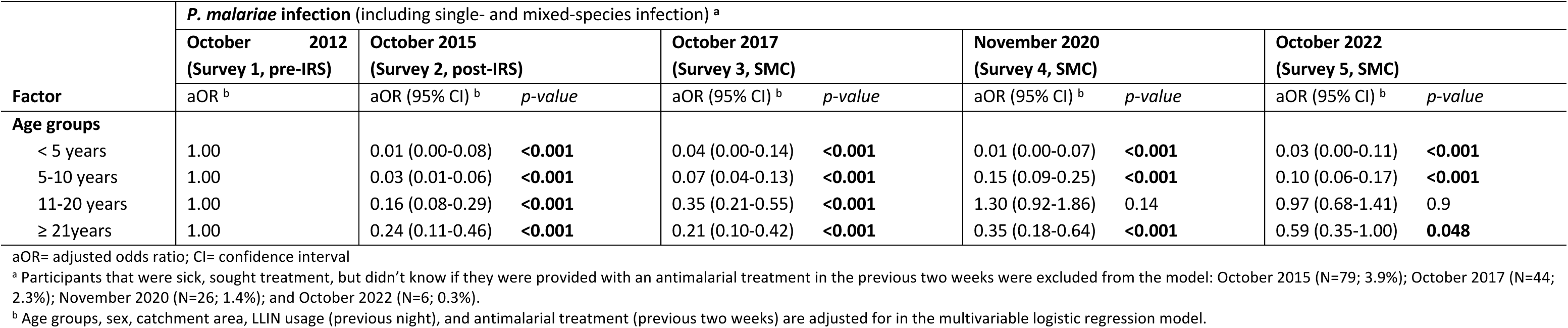
Stratum-specific estimates for the association between age groups and *P. malariae* prevalence during each study time point.

**S4 Table.**
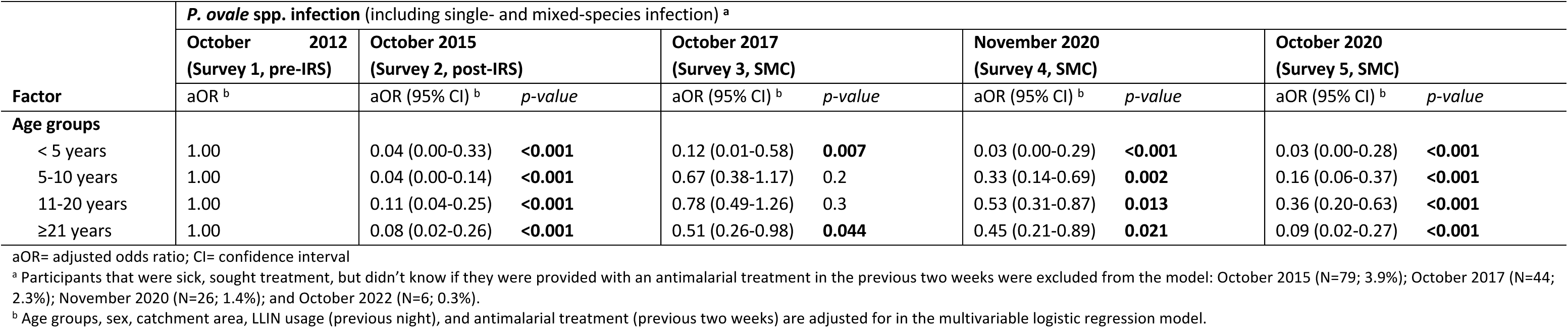
Stratum-specific estimates for the association between age and *P. ovale* spp. prevalence during each study time point.

**Fig S1.**
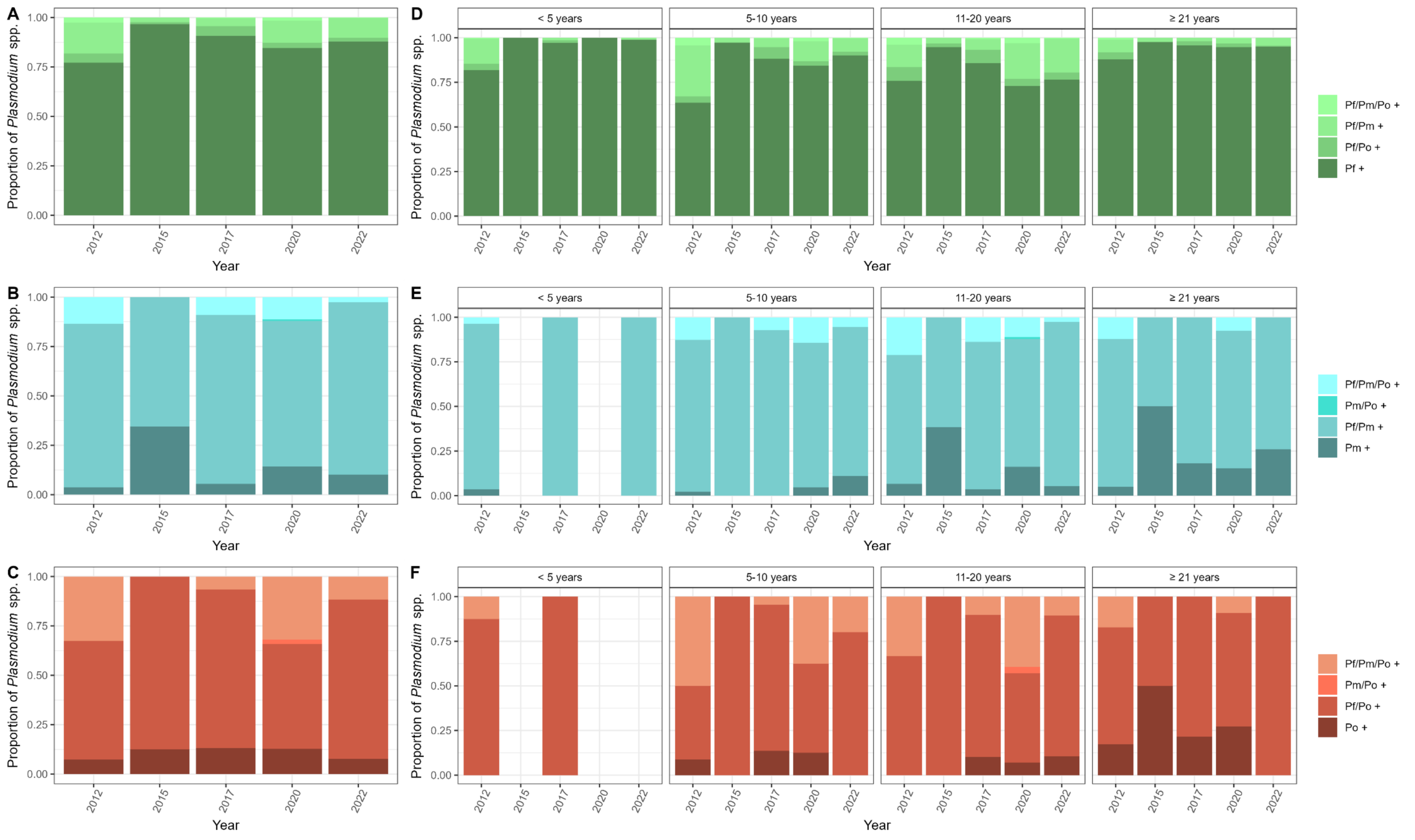
Proportion of single- and mixed-*Plasmodium* spp. infections identified using the species-specific *18S rRNA* PCR during each study time point. Proportion of single- and mixed-species infections (**A-C**) in the population and (**D-F**) by age group for *P. falciparum* (green)*, P. malariae* (blue), and *P. ovale* spp. (red) during each study time point. For each *Plasmodium* spp. darker shades represent those isolates with single-species infections, while the lighter shades denote those isolates with mixed-species infection (including double- and triple-species infections). Blank or white spaces indicate no infections detected by the species-specific *18S rRNA* PCR.

